# Unprecedented disruptions of lives and work – a survey of the health, distress and life satisfaction of working adults in China one month into the COVID-19 outbreak

**DOI:** 10.1101/2020.03.13.20034496

**Authors:** Stephen X Zhang, Yifei Wang, Andreas Rauch, Feng Wei

## Abstract

What are the health and wellbeing of people during the COVID-19 outbreak? We aim to assess the health and wellbeing of normal adults living and working after one month of confinement to contain the COVID-19 outbreak in China. On Feb 20–21, 2020, we surveyed 369 adults in 64 cities in China that varied in their rates of confirmed coronavirus cases on their health conditions, distress and life satisfaction. The participants also reported their work status, whether they had chronic health issues, and the number of hours they exercised per day in the past week. 27% of the participants worked at the office, 38% resorted to working from home, and 25% stopped working due to the outbreak. Those who stopped working reported worse mental and physical health conditions as well as distress. The severity of COVID-19 in an individual’s home city predicts their life satisfaction, and this relationship is contingent upon individuals’ existing chronic health issues and their hours of exercise. Our evidence supports the need to pay attention to the health of people who were not affected by the virus epidemiologically, especially for people who stopped working during the outbreak. Our results highlight that physically active people might be more susceptible to wellbeing issues during the lockdown. Policymakers who are considering introducing restrictive measures to contain COVID-19 may benefit from understanding such health and wellbeing implications.

## INTRODUCTION

Since the escalation of COVID-19 to a public health emergency in China on Jan 21, 2020, over a billion people across China have faced restrictions due to varying degrees of confinement such as banning public transport, restricting movement, and imposing a 14-day quarantine after travel (Li et al., 2020; Wang et al., 2020). One month into the outbreak, many people were still not working or exercising as usual, which may have associated implications on health and wellbeing. Yet, the implications of the unusually prolonged state of not working and exercising on individuals’ health and wellbeing remain unknown. It is important to understand not only the implications of the restrictions on COVID-19 disease prevalence rates but also the implications of such unprecedented disruptions on the health and wellbeing of the community (Brooks et al., 2020). From a policy perspective, understanding the health and wellbeing of people under the varying degrees of lockdown in China has implications for countries that are just starting to fight coronavirus, as such restrictions started in Korea, Italy, parts of the US, etc. in March (Moodie Davitt Report, 2020).

To control the COVID-19 outbreak, China has enacted restrictive measures “unprecedented in public health history” as stated by WHO (Reuters, 2020). On January 23, 2020, China locked down Wuhan, a metropolitan area of 12 million people (ABC News, 2020). The lockdown in Wuhan soon triggered similar measures in all 15 other cities in Hubei province with a total of 57 million people. Other prefectures (the administrative areas of a city) in China subsequently implemented varying levels of restrictive measures. For instance, the prefecture of Wenzhou in Zhejiang province restricted its citizens in such a way that only one person per household could leave home once every two days. Such restrictive measures in China seemed to be effective in containing the spread of COVID-19 by mid-February and were applauded by WHO (The Washington Post, 2020). However, those measures have disrupted people’s jobs and lives immensely and hence may have important implications for their health and wellbeing.[8] For example, evidence from the SARS crisis indicated that reduced mobility affected the wellbeing of quarantined residents in a complex manner. An article in *Lancet COVID-19 series* specifically highlighted that people who had been in quarantine reported a high prevalence of symptoms of psychological distress and disorder and some of these symptoms seem to stay long after the quarantine (Lima et al., 2020). A paper in psychiatry research highlighted the need to study mental health in Iran (Zandifar and Badrfam, 2020). These papers are critical because across the world people who did not carry the virus epidemiologically but had their work and life disrupted to varying degrees (Duan and Zhu, 2020; Xiang et al., 2020; Bao et al., 2020). In this article, we aim to use existing scales of health, distress and life satisfaction to identify the health and wellbeing of people one month into the disruption caused by confinement measures to contain COVID-19 outbreak by their work status, chronic health conditions, and exercising hours.

Understanding the health and wellbeing implications of the measures introduced to reduce the COVID-19 infection allows better-informed decisions. As many parts of the world are starting to consider measures to contain COVID-19, with South Korea and Italy having introduced lockdowns in early March 2020 (Cohen and Kupferschmidt, 2020), we may benefit from understanding the health and wellbeing implications of the measures implemented early on in China.

## METHODS

We conducted a cross-sectional survey one month into the COVID-19 outbreak on February 20–21, 2020, about one month into the COVID-19 emergency in China. All the participants were adults not affected by the virus epidemiologically but they lived in locations that were affected by COVID-19 to varying degrees. To cover people in areas of varying severity of COVID-19, we surveyed adults from 64 prefectures across China. The 64 prefectures were chosen to cover a wide spectrum of regions based on the severity of COVID-19 and should not be taken as a representative national sample. All respondents agreed to participate in the study, which was approved by the ethics committee at Tongji University (#20200211). We reached 529 adults, and 369 of them answered the survey, with a response rate of 69.8%. The participants were not involved in the design, or conduct, or reporting, or dissemination plans of this research.

We assessed individual health by the Short Form-12 (SF12), a standard scale on mental and physical health function (Ware et al., 1996). The scale had been translated into Chinese and validated in China (Zhang et al., 2011). SF12 contains 12 items and 8 dimensions: physical functioning (2 items), role physical (2 items), bodily pain (1 item), general health (1 item), vitality (1 item), social functioning (1 item), role emotional (2 items) and mental health (2 items). The eight dimensions form two composite scores of physical and mental composite scale (PCS and MCS), with a possible score ranging from 0 to 100 (Ware et al., 2006). As a formative score, a higher SF12 score indicates a better health condition.

We measured distress by the six-item Kessler psychological distress scale (K6) with a Cronbach’s alpha of 0.79 (Kessler et al., 2002). We measured life satisfaction with the Satisfaction With Life Scale (SWLS) (Diener et al., 1985), which consists of five items with a Cronbach’s alpha of 0.72. The adults also provided their socio-demographic characteristics, such as gender, age, education, and their location (prefecture). Using their locations, we searched for the number of confirmed COVID-19 cases in their prefectures on February 20 as well as the total population to calculate the number of confirmed cases per 10,000 people as an indicator of the severity of COVID-19 at their location. The number of cases per 10,000 people (i.e. infected rate) varied from 42.45 (Wuhan) to 0.01 on February 21 (National Health Commission of the PRC, 2020).

Because COVID-19 is more dangerous for people with comorbidity (Gates, 2020), it is likely that people who have ongoing medical issues would suffer more during the outbreak and therefore we asked whether the participants had any chronic disease. On the contrary, people who lead a healthy lifestyle and exercise often would be expected to fare better during the outbreak. Hence, we also asked the participants to indicate ‘how many hours did you exercise per day during the past week’. The restrictive measures of COVID-19 also caused major disruption to people’s work. By the time of our survey on February 20, the growth rate of COVID-19 cases in China had fallen to single percentages per day. Some people still stopped work, while some had returned to work in offices, and others were working at home. All individuals reported their work status.

We report the descriptive statistics of the study variables and the regression models to examine the relationships. The first and second author did the analyses on unweighted data with STATA 16.0, and statistical significance was assessed by p□<□0.05.

## RESULTS

Table 1 presents the descriptive characteristics of the participants. From January 21 to February 20, in the one month of the restrictive measures, 124 (33.6%) of the participants had not left home at all, 51 (13.8%) had left their home only once, and 81 (22.0%) had left their home more than five times. At the time of the survey, 99 (26.8%) were going to work at their office; 93 (25.2%) had stopped working; and 139 (37.7%) resorted to working from home; 32 (8.7%) participants had not been working before the outbreak started; and 6 (1.6%) reported losing their work during the COVID-19 outbreak.

**Table 1.** Descriptions of the participants (n=369)

| Variable | Count or mean | Percentage |
| --- | --- | --- |
| Gender |  |  |
| male | 204 | 55.0% |
| female | 165 | 45.0% |
| Age (years) Mean (SD) | 36.6 (10.5) |  |
| Education level |  |  |
| secondary school or below | 40 | 10.8% |
| high school or vocational school | 43 | 11.7% |
| two-year college degree | 57 | 15.5% |
| bachelor degree | 157 | 42.5% |
| postgraduate degree | 72 | 19.5% |
| Marital status |  |  |
| single | 105 | 28.5% |
| married | 252 | 68.3% |
| others | 12 | 3.2% |
| Chronic disease |  |  |
| yes | 45 | 12.2% |
| no | 324 | 87.8% |
| Job status |  |  |
| worked at office | 99 | 26.8% |
| worked at home | 139 | 37.7% |
| stopped working | 93 | 25.2% |
| no work before and during outbreak | 32 | 8.7% |
| lost job during the outbreak | 6 | 1.6% |
| Total number of times out of home in the last month |  |  |
| never | 124 | 33.6% |
| once | 51 | 13.8% |
| twice to five times | 113 | 30.6% |
| more than five times | 81 | 22.0% |
| Exercise hours per day in the past week |  |  |
| 0 | 51 | 13.8% |
| 0~1 | 226 | 61.2% |
| 1~2.5 | 63 | 17.1% |
| ≥2.5 | 29 | 7.9% |
| Health condition |  |  |
| physical composite scale: Mean (SD) | 49.55 (7.00) |  |
| mental composite scale: Mean (SD) | 48.74 (9.30) |  |
| Distress: Mean (SD) | 1.41 (0.46) |  |
| Life Satisfaction: Mean (SD) | 3.22 (0.64) |  |

Note: The scores of physical composite scale range from 27.54 to 63.83; the scores of mental composite scale range from 18.83 to 68.06; the scores of distress and life satisfaction both range from 1 to 5.

In terms of exercise, 51 (13.8%) people had not exercised at all during the past week; 226 (61.2%) exercised but for less than 1 hour per day; 63 (17.1%) exercised 1 to 2.5 hours per day; and 29 (7.9%) exercised more than 2.5 hours per day. Of the participants, 45 (12.2%) had chronic diseases.

Based on the scoring algorithm of SF12,^16^ the participants scored 48.74 (SD 9.30) in mental health (MCS) and 49.55 (SD 7.00) in physical health (PCS). The mean values of distress and life satisfaction of the participants were 1.41 (SD 0.46) and 3.22 (SD 0.64) respectively.

### People who worked in the office, worked at home, or had stopped working differed in health (SF12) and distress (K6)

Table 2 shows the results of regressing SF12 on the job status of the participants one month into the COVID-19 outbreak. Compared with people who stopped working during the outbreak, people who worked at their office had better mental health (β=3.46, p=0.01, 95% CI 0.79 to 6.14) and physical health (β=2.19, p=0.01, 95% CI 0.20 to 4.18). Moreover, people who worked at home also had better mental health than those who stopped working (β=2.60, p=0.03, 95% CI 0.45 to 5.16).

**Table 2.**
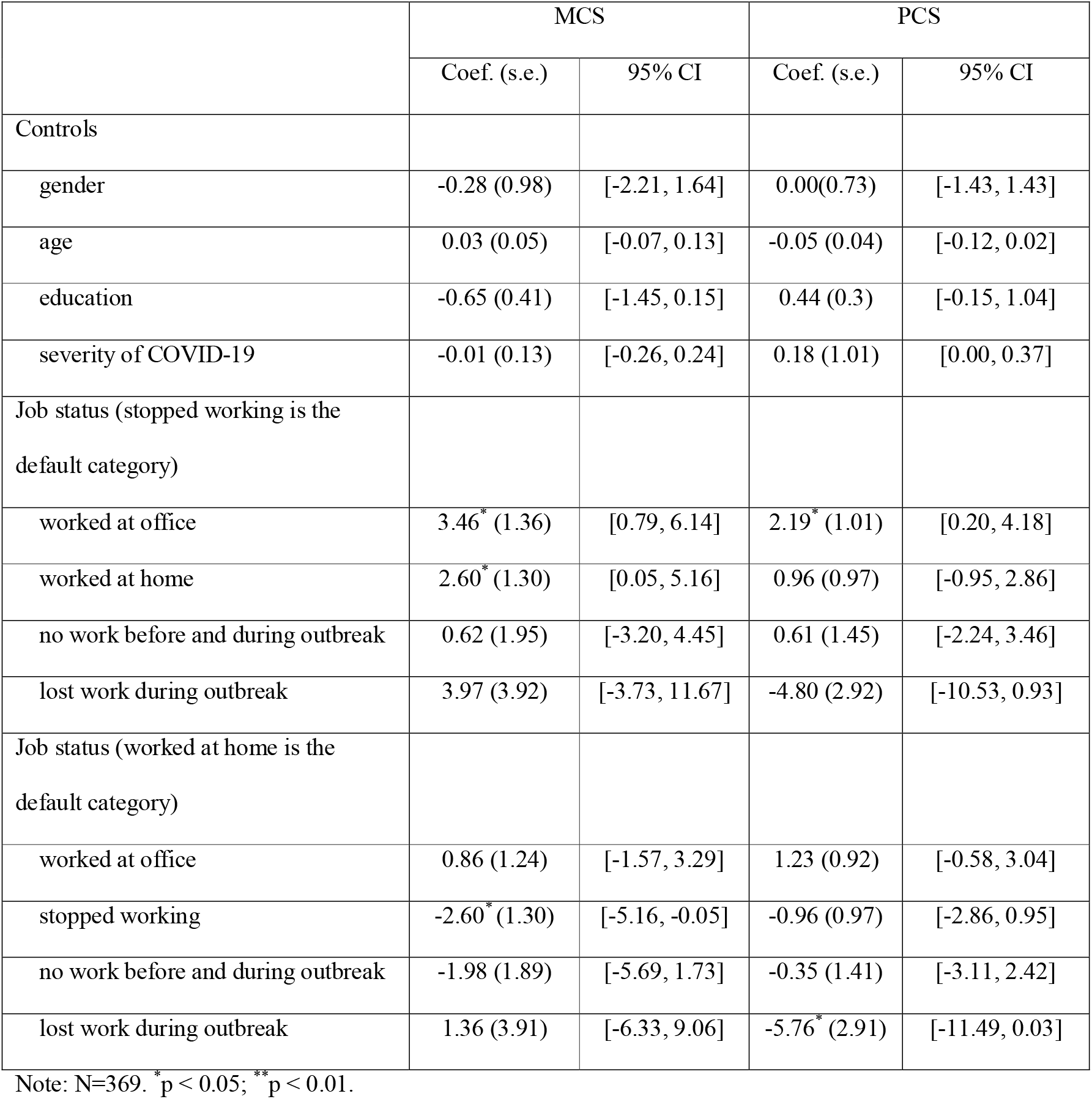
OLS regression results on the Mental Composite Scale (MCS) and Physical Composite Scale (PCS) of SF12 by job status.

|  | MCS |  | PCS |  |
| --- | --- | --- | --- | --- |
|  | Coef. (s.e.) | 95% CI | Coef. (s.e.) | 95% CI |
| Controls |  |  |  |  |
| gender | -0.28 (0.98) | [-2.21, 1.64] | 0.00(0.73) | [-1.43, 1.43] |
| age | 0.03 (0.05) | [-0.07, 0.13] | -0.05 (0.04) | [-0.12, 0.02] |
| education | -0.65 (0.41) | [-1.45, 0.15] | 0.44 (0.3) | [-0.15, 1.04] |
| severity of COVID-19 | -0.01 (0.13) | [-0.26, 0.24] | 0.18 (1.01) | [0.00, 0.37] |
| Job status (stopped working is the default category) |  |  |  |  |
| worked at office | 3.46* (1.36) | [0.79, 6.14] | 2.19* (1.01) | [0.20, 4.18] |
| worked at home | 2.60* (1.30) | [0.05, 5.16] | 0.96 (0.97) | [-0.95, 2.86] |
| no work before and during outbreak | 0.62 (1.95) | [-3.20, 4.45] | 0.61 (1.45) | [-2.24, 3.46] |
| lost work during outbreak | 3.97 (3.92) | [-3.73, 11.67] | -4.80 (2.92) | [-10.53, 0.93] |
| Job status (worked at home is the default category) |  |  |  |  |
| worked at office | 0.86 (1.24) | [-1.57, 3.29] | 1.23 (0.92) | [-0.58, 3.04] |
| stopped working | -2.60* (1.30) | [-5.16, -0.05] | -0.96 (0.97) | [-2.86, 0.95] |
| no work before and during outbreak | -1.98 (1.89) | [-5.69, 1.73] | -0.35 (1.41) | [-3.11, 2.42] |
| lost work during outbreak | 1.36 (3.91) | [-6.33, 9.06] | -5.76* (2.91) | [-11.49, 0.03] |

Note: N=369. \* $p < 0.05$ ; \*\* $p < 0.01$ .

We did further analysis on the eight specific dimensions of SF12. Compared with people who returned to work at their office, those who stopped working reported lower general health (β=-0.28, p=0.03, 95% CI −0.55 to −0.03), mental health (β=-0.38, p=0.002, 95% CI −0.63 to −0.14), and increased limitations for physical issues (β=-0.16, p=0.009, 95% CI −0.28 to −0.04) and emotional issues (β=-0.13, p=0.03, 95% CI −0.26 to −0.01). There were fewer differences between people who worked at home and worked at offices, except those who worked at home reported more limitations for physical issues than those who worked at offices (β=-0.13, p=0.02, 95% CI −0.24 to −0.02).

There were also certain dimensions of SF12 that did not vary much by job status. For instance, people who stopped working and those who worked at their offices did not differ in bodily pain (β=0.16, p=0.16, 95% CI −0.06 to 0.39), physical function (β=0.10, p=0.14, 95% CI −0.03 to 0.24), and social function (β=0.22, p=0.26, 95% CI −0.16 to 0.59). Those who stopped working and those who worked at home respectively also did not differ in the same dimensions of bodily pain (β=0.15, p=0.19, 95% CI −0.07 to 0.36), physical function (β=0.00, p=0.96, 95% CI −0.13 to 0.13), and social function (β=0.19, p=0.29, 95% CI −0.17 to 0.55).

Similar findings emerged from the regression results on distress (K6) and life satisfaction (Table 3). People who worked at their office suffered less distress than people who stopped working (β=-0.13, p=0.05, 95% CI −0.25 to 0.00). Also, people who worked at their office reported higher life satisfaction than those who stopped working (β=0.23, p=0.02, 95% CI 0.03 to 0.43).

**Table 3.** OLS regression results on distress and life satisfaction by job status.

|  | Distress (K6) |  | Life satisfaction |  |
| --- | --- | --- | --- | --- |
|  | Coef. (s.e.) | 95% CI | Coef. (s.e.) | 95% CI |
| Controls |  |  |  |  |
| gender | 0.03 (0.05) | [-0.06, 0.12] | 0.25** (0.05) | [0.11, 0.39] |
| age | -0.00 (0.00) | [-0.01, 0.00] | 0.02*** (0.00) | [0.01, 0.03] |
| education | 0.05** (0.02) | [0.01, 0.08] | 0.03 (0.03) | [-0.03, 0.09] |
| severity of COVID-19 | -0.00 (0.01) | [-0.01, 0.01] | -0.01 (0.01) | [-0.02, 0.01] |
| Job status (stopped working is the default category) |  |  |  |  |
| worked at office | -0.13* (0.06) | [-0.25, 0.00] | 0.23* (0.10) | [0.03, 0.43] |
| worked at home | -0.06 (0.06) | [-0.18, 0.06] | 0.06 (0.10) | [-0.14, 0.25] |
| no work before and during outbreak | -0.15 (0.10) | [-0.34, 0.04] | 0.15 (0.15) | [-0.14, 0.44] |
| lost work during outbreak | 0.22 (0.20) | [-0.17, 0.61] | 0.02 (0.29) | [-0.56, 0.59] |
| Job status (work at home is the default category) |  |  |  |  |
| worked at office | -0.07 (0.06) | [-0.18, 0.04] | 0.17 (0.09) | [-0.01, 0.36] |
| stopped working | 0.06 (0.06) | [-0.06, 0.28] | -0.06 (0.09) | [-0.25, 0.14] |
| no work before and during outbreak | -0.09 (0.09) | [-0.27, 0.09] | 0.09 (0.14) | [-0.18, 0.37] |
| lost work during outbreak | 0.28 (0.20) | [-0.11, 0.66] | -0.04 (0.29) | [-0.61, 0.54] |

Note: N=369. \* $p < 0.05$ ; \*\* $p < 0.01$ .

### Are people in areas more affected by COVID-19 less satisfied with their life? It depends

Next, we analyzed how the severity of COVID-19 in individual locations predicts individuals’ life satisfaction. Table 4 shows that the relationship between the severity of COVID-19 and individual life satisfaction depends on individuals’ existing health and exercise status. The severity of COVID-19 had a negative relationship with the life satisfaction only for people with chronic medical issues (β=-2.11, p=0.04, 95% CI −4.09 to −0.13) but not for people without chronic medical issues (β=-0.01, p=0.51, 95% CI −0.02 to 0.01). We plot the effect of the severity of COVID-19 on life satisfaction by whether the individuals had chronic medical issues in Figure 1.

**Table 4.** The severity of COVID-19 in a location interacts with individuals’ chronic health condition and exercise time to predict their life satisfaction.

|  | Life satisfaction |  |
| --- | --- | --- |
|  | Coef. (s.e.) | 95% CI |
| Controls |  |  |
| gender | 0.25*** (0.07) | [0.10, 0.39] |
| age | 0.02*** (0.00) | [0.01, 0.03] |
| education | 0.04 (0.03) | [-0.01, 0.11] |
| Job status (worked at office is the default category) |  |  |
| worked at home | 0.07 (0.06) | [-0.04, 0.18] |
| stopped working | 0.13* (0.06) | [0.00, 0.25] |
| no work before and during outbreak | -0.03 (0.09) | [-0.21, 0.16] |
| lost work during outbreak | 0.34 (0.20) | [-0.04, 0.73] |
| Severity of COVID-19 | -2.04* (0.99) | [-4.00, -0.08] |
| Chronic health condition<sup>1</sup> | -0.20 (0.21) | [-0.61, 0.20] |
| Exercise hour | 0.04 (0.03) | [-0.01, 0.09] |
| Interaction |  |  |
| severity of COVID-19 × chronic health condition | 2.08* (0.99) | [0.13, 4.04] |
| severity of COVID-19 × exercise hours | -0.02** (0.01) | [-0.04, -0.01] |

Note: N=369. \*p < 0.05; \*\*p < 0.01. 1 = people with chronic disease, 2 = people without chronic disease.

**Figure 1:**
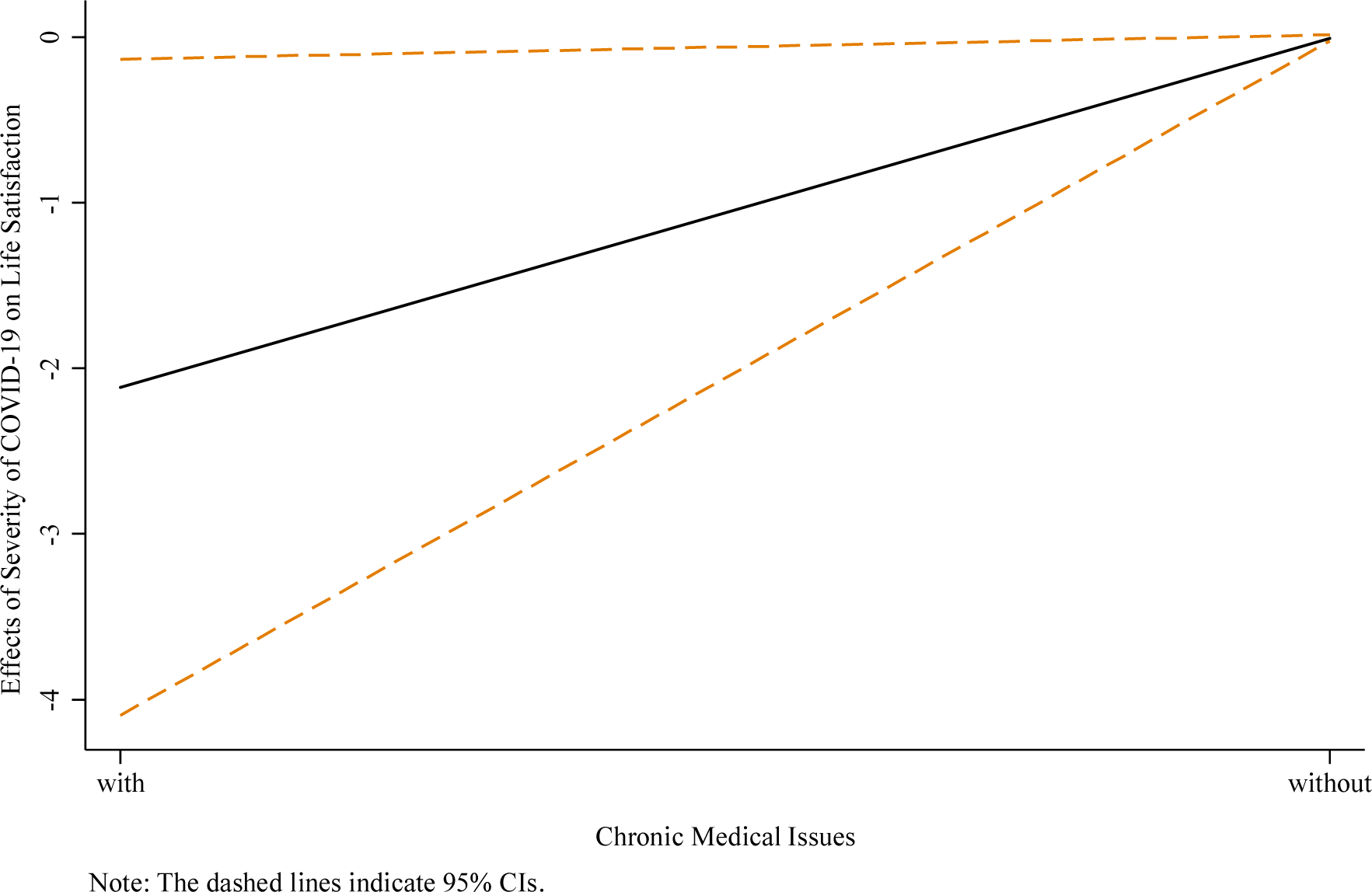
The effect of the severity of COVID-19 on life satisfaction depends on the chronic medical issues of the individuals.

The results also indicate that the relationship between the severity of COVID-19 in a location and life satisfaction depends on individuals’ level of exercise, but in a direction opposite to our expectation. While we expected people who exercised more during the outbreak had a healthy lifestyle and would be less influenced, for people who exercised more than 2.5 hours per day during the outbreak, the relationship between the severity of COVID-19 in their location and life satisfaction was significantly negative (e.g. for people who exercised 3 hours per day: β=-0.02, p=0.04, 95% CI −0.04 to 0.00). The relationship is not significant for people who exercised between 1 and 2.5 hours a day during the outbreak. Surprisingly, for people who exercised 0.5 hours or less per day during the outbreak, their life satisfaction was significantly positively associated with more affected locations (e.g. for people who did not exercise: β=0.04, p=0.02, 95% CI 0.01 to 0.08). We plot the effect of the severity of COVID-19 in the location on life satisfaction by the exercise hours of individuals in Figure 2.

**Figure 2:**
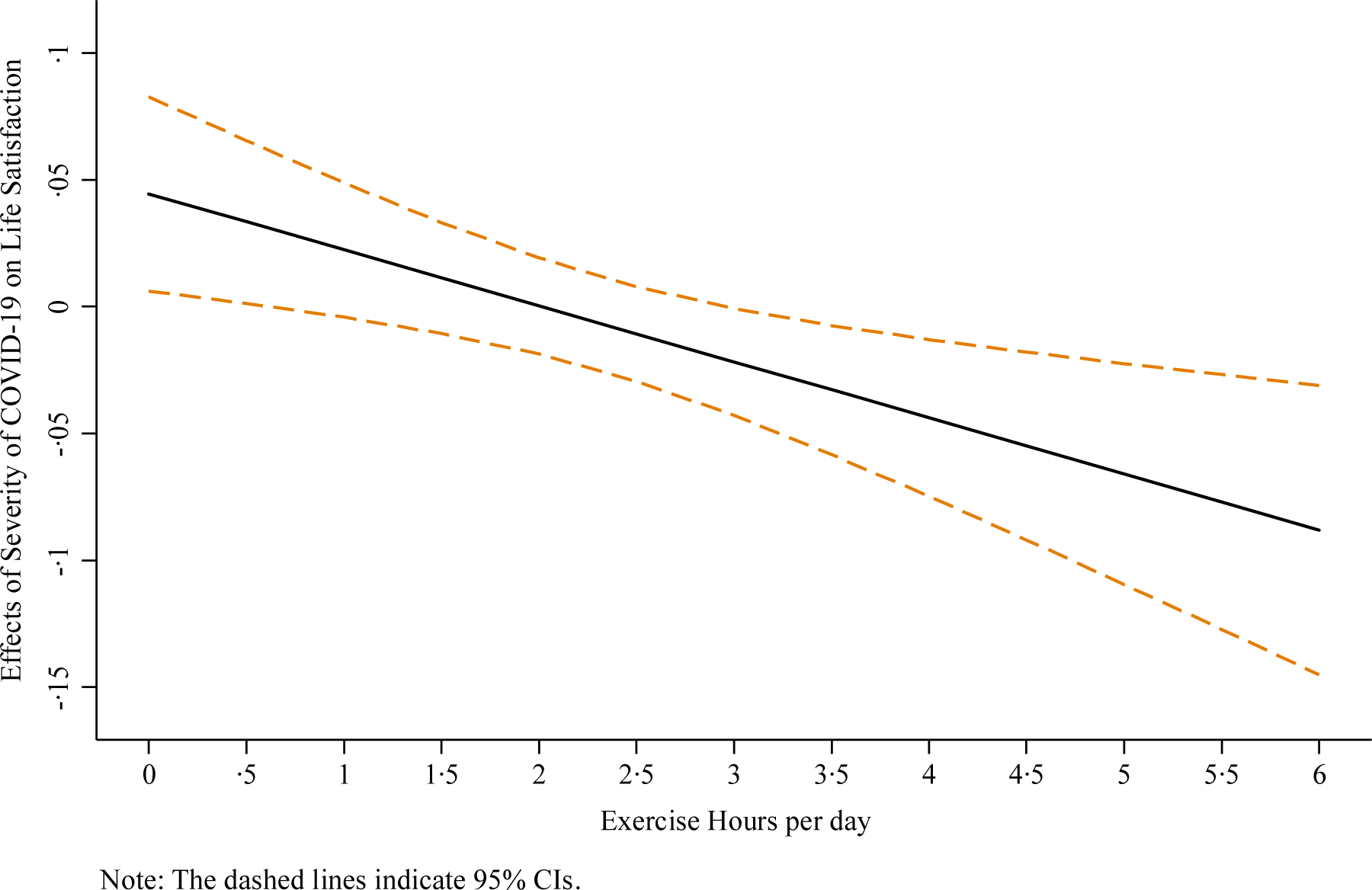
The effect of the severity of COVID-19 on life satisfaction depends on the exercising hours of the individuals.

## DISCUSSION

Although the hardline restrictive measures in China showed success in containing COVID-19 after a month, there is little research on the extent to which the disruptions affected people in the community. Our findings show that adults who were not working reported worse health, as captured by SF12 in certain dimensions as well as distress (K6). The insignificant differences in the dimensions of SF12 in bodily pain, physical function and social function are expected, because people who were not affected by the virus directly would not differ much in those dimensions.

The results on life satisfaction are more nuanced. Overall, the severity of COVID-19 in a prefecture did negatively predict people’s life satisfaction in that prefecture, with this relationship depending on individuals’ existing medical conditions and exercise levels. For individuals who exercised a lot during the outbreak (>2.5 hours per day), life satisfaction was significantly negatively associated with more affected locations; on the contrary, for individuals who exercised half an hour a day or less, life satisfaction was significantly positively associated with more affected locations. Maybe these people could better justify or rationalize their inactive lifestyles in more severely affected cities. The finding that those who exercised a lot (>2.5 hours per day) were less satisfied in more affected cities suggests we may need to pay attention to more physically active individuals, who might be more frustrated by the restrictions due to the outbreak.

The study has certain limitations. First, this study relied on an observational survey. Because the measures of the dependent variables of SF12, distress and life satisfaction used Likert scales, we tried to use predictors that were non-Likert scales, such as job status and severity of COVID-19 calculated based on archival data using the reported locations. Second, our sample is not a national representative sample. Our focus was to examine the differential effects on adults depending on the level of disruption, as captured by their job status, existing chronic issues, and exercise levels to identify who in the community of non-COVID-19 cases might need the most help for policymakers and potential caregivers. Third, even though we had data from people who had not been working even before the outbreak started (8.7%) and people who went out of work during the outbreak (1.6%), the sample size of those two groups was small and we are cautious not to report them in the findings. Nevertheless, people with these two job statuses could be important targets for future studies. We provide preliminary evidence on the health conditions of adults in COVID-19 affected regions. The identification of who might be more affected by COVID-19, not epidemiologically but simply by working and living in affected regions, carries important implications. Such identification can help to prioritize those who might need more help, and psychologists, mental health professionals and social workers can provide services to start addressing at least the mental health issues, even during the lockdown. Affected regions are growing globally by the day, and COVID-19 is no longer confined to China (Cohen and Kupferschmidt, 2020; Croakey, 2020). To contain COVID-19 transmission (Li et al., 2020), policymakers in other countries are considering implementing restrictive measures (Kupferschmidt and Cohen, 2020). We present this early evidence of disruptions one month into the outbreak to provide evidence on the health of the general community living and working under restrictive measures.

## Data Availability

upon request

## Contributors

Stephen X. Zhang contributed to the planning, conduct, and reporting of the work and is responsible for the overall content as the guarantor. Yifei Wang cleaned the data, made the figures and tables, and prepared the submission files. Andreas Rauch helped with the writing of the paper. Feng Wei led the data collection and obtained necessary funding and ethical approval. The corresponding author attests that all listed authors meet authorship criteria and that no others meeting the criteria have been omitted.

